# Reported case fatality as a target and trial metric in endemic zoonoses: a framework and ecological analysis of Lassa fever surveillance in Nigeria, 2017–2026

**DOI:** 10.64898/2026.10.14.26365510

**Authors:** David Simons

## Abstract

**Introduction:** Reported case fatality ratios (CFRs) are used to set national targets, such as Nigeria’s goal of a Lassa fever CFR below 10%, and to plan vaccine and therapeutic trials, yet they also move with testing, death reporting and where cases occur. Using the rise in Nigeria’s reported Lassa fever CFR from 18.9% at week 23 of 2025 to 25.0% in 2026 as a test, I evaluated what published data can show.

**Methods:** This ecological study extracted weekly state-level counts from 474 Nigeria Centre for Disease Control and Prevention situation reports (2017-2026). Following a pre-specified plan, years were compared at phase-matched points, year-on-year changes were decomposed into within-state and between-state components, and the ascertainment change needed to explain 2026 was calculated. Comparing the published cumulative and summed weekly series was added after the plan was fixed.

**Results:** By week 36 of 2026, 70 deaths had been added to the cumulative totals outside any week’s figures (4 in all of 2025), so the published 2026 CFR exceeded 2025 by 5.4 to 6.2 percentage points at every comparison point. On the weekly series, 2026 did not differ from 2025 at the phase-matched midpoint (+0.3 percentage points, 95% CI -5.0 to 7.6), and within-state (-1.1, 95% CI -4.7 to 3.0) and between-state (+0.5, -2.0 to 2.9) components spanned zero. An additional 279 unconfirmed survivors (6% of suspected cases not confirmed) would bring the published 2026 CFR down to the 2025 level.

**Conclusion:** The 2026 rise came from deaths added to cumulative totals, not the weekly counts, and cannot be attributed to care. Reported CFRs alone cannot track national targets or set trial baselines. Data already collected in case-based surveillance (specimens tested, sampling at or after death, outcome ascertainment, late-added deaths by week of confirmation) could be published to close this gap.

**Key messages:** *What is already known on this topic.:* Reported case fatality ratios (CFRs) are used to set targets and plan trials for endemic zoonoses such as Lassa fever, but methods for separating a change in reported CFR from changes in testing and reporting were developed for epidemics spread person to person.

*What this study adds.:* In Nigeria, the published 2026 rise in Lassa fever CFR came from deaths added to cumulative totals outside any week’s figures; on the weekly figures it did not differ from 2025, and the published reports allow only 2 of 11 mechanisms that move a reported CFR to be evaluated fully.

*How this study might affect research, practice or policy.:* A minimum reporting set deliverable from data already collected in case-based surveillance (specimens tested, the share sampled at or after death, the share with a known outcome and late-added deaths by week of confirmation) would let national CFR targets be tracked and give trialists and funders baselines that separate surveillance from severity.

## Introduction

The case fatality ratio (CFR, reported in surveillance as the case fatality rate) is frequently treated as the primary measure of pathogen virulence and clinical severity. It shapes how an outbreak is perceived and how diseases are prioritised, and where research, countermeasure investment, and clinical resources are directed [1]. Reported CFRs also serve as targets, against which national programmes judge progress, and as inputs to the design of vaccine and therapeutic trials. A reported CFR is the share of confirmed cases who died, where both numerator and denominator are produced by surveillance. It is not an intrinsic biological property of a pathogen. It fluctuates with who is tested, which deaths are found and recorded, where cases occur and when the value is calculated, as well as with the risk of death [2,3].

Methods for these surveillance biases were developed mainly for newly emerging epidemics with sustained human-to-human transmission [2], and applied recently to COVID-19 [4]. In those settings, deaths lag notifications, and these biases attenuate as an epidemic progresses and testing expands [2,3]. Endemic zoonoses differ. Human cases arise from repeated, independent spillover from wildlife reservoirs across dispersed geographies [5]. Detection is dependent on clinical suspicion during acute presentations of undifferentiated febrile illness, often against a high background of other causes of fever, such as malaria [6,7]. Because transmission chains are short, surveillance figures are not modelled as expanding epidemic waves; instead counts are compared year on year, with each transmission season representing a distinct configuration of spillover pressure, diagnostic practice, and reporting behaviour, and detection can depend on the outcome itself [8]. A review of the literature (appendix) found 12 studies that formally separated a change in a reported CFR from changes in testing or reporting, all of them in diseases spread from person to person, 11 in COVID-19. The tools for reading a change in reported CFR have therefore been built for epidemics, not for zoonoses whose figures are compared from one season to the next.

Lassa fever exemplifies these structural challenges. It is endemic across West Africa, and in Nigeria the Nigeria Centre for Disease Control and Prevention (NCDC) has published weekly situation reports since January 2017, the main public record of its seasonal outbreaks [9]. Each report tracks the CFR among laboratory-confirmed cases against the same epidemiological week of the previous year. Most infections with *Mammarenavirus lassaense* (LASV; Arenaviridae) are considered to be asymptomatic or mild [10], so confirmed cases are a small, severity-selected subset of infections [11]. Annual CFRs from the weekly reports have ranged from 12.6% (2020) to 25.0% (2018) since 2018. Published estimates vary so widely that a meta-analysis attributed almost all variation between studies to heterogeneity (I² 95%) [12]. In a community cohort with active case-finding, five of 39 people with Lassa fever died (13%), although cases there were found and treated early [7]. Cases are reported by state, diagnostic capacity has expanded unevenly, and some treatment centres take referrals from neighbouring states, so a state’s count can reflect where patients are treated as well as where they were infected [13]. Nigeria’s national strategy sets a target of a Lassa fever CFR below 10%, measured with the published cumulative figure [14], and the WHO target product profile for Lassa fever vaccines notes that the true incidence is unknown and that existing estimates are biased by limited testing capacity [15].

The 2026 season provides a test of these uses. At week 23 the reported cumulative CFR was 25.0%, compared with 18.9% in the same period of 2025. This rise was read as evidence that patients were presenting late along the care pathway, with calls for earlier diagnosis and faster referral, although the authors noted that a CFR among confirmed cases cannot by itself separate a change in the risk of death from a change in testing [16].

Published situation reports make surveillance data public week by week and support the response, research and trial design, but they are built for situational awareness: they report counts, not the intervals, denominators and case characteristics that would separate a change in ascertainment from a change in outcome.

Using Nigerian Lassa fever surveillance as a case study, I evaluated what public surveillance data can demonstrate about changes in a reported CFR in four stages: establishing a framework of 11 mechanisms that can shift a reported CFR and the data signature each would leave; auditing published situation reports to determine which signatures can be resolved; evaluating the resolvable mechanisms in each year-on-year transition from 2018 to 2026, following a pre-specified analysis plan; and investigating a discrepancy between weekly counts and cumulative totals to identify the driver of the 2026 rise and specify the minimum reporting dataset required for future evaluation.

## Methods

### Data

In this ecological analysis of published surveillance reports, I extracted weekly counts of suspected, confirmed and probable cases and of deaths among confirmed cases, by state, from NCDC Lassa fever situation reports from January 2017 to 2026 week 36 (the week ending 6 September), the last report published before the data freeze of 1 October 2026. Only the published reports were used; the underlying case records were not available. Every value was manually extracted from the report [17] and checked against the totals printed in it; 80 of 18,152 values (0.4%) had been mistyped during extraction, transcription errors rather than errors in the reports, and identified errors were corrected (appendix). Deaths reported only as a national total, in seven reports, were allocated to states in proportion to their confirmed cases that week rather than dropped, which would have lowered those years’ CFRs, and redrawn in every bootstrap replicate. Rules for missing weeks and late reports are in the appendix. 2017 is descriptive only, because 18 of its weeks have no report; 2020 and 2021 are included without adjustment for COVID-19 disruptions. Ethical approval was not sought, as the data are publicly available and aggregated. The analyses followed a pre-specified plan, fixed on 4 October 2026 before they were run on the final data (see Data availability statement); deviations from it, including corrections to the extract made after it was fixed, are listed in the appendix.

### Definitions

Year means epidemiological year as numbered in the reports. Transmission peaks from about December to April, so one transmission season straddles calendar years. Reported CFR is deaths among confirmed cases divided by confirmed cases, per NCDC. The proportion of suspected cases confirmed is a proxy, not test positivity, because it includes the decision to test a suspected case, which is itself one of the mechanisms under study. The analyses use the weekly figures, the only series that records each case and death in the week it was reported; the cumulative totals also take in deaths reported later, in amounts that vary between years, so only the weekly series allows years to be compared at the same point in the season (the two series are compared in A9).

### Mechanisms and evaluability

I listed the mechanisms that can move a reported CFR, the signature each would leave in the published indicators, the marker that would identify it, and whether the published reports contain that marker (Table 1).

**Table 1:**
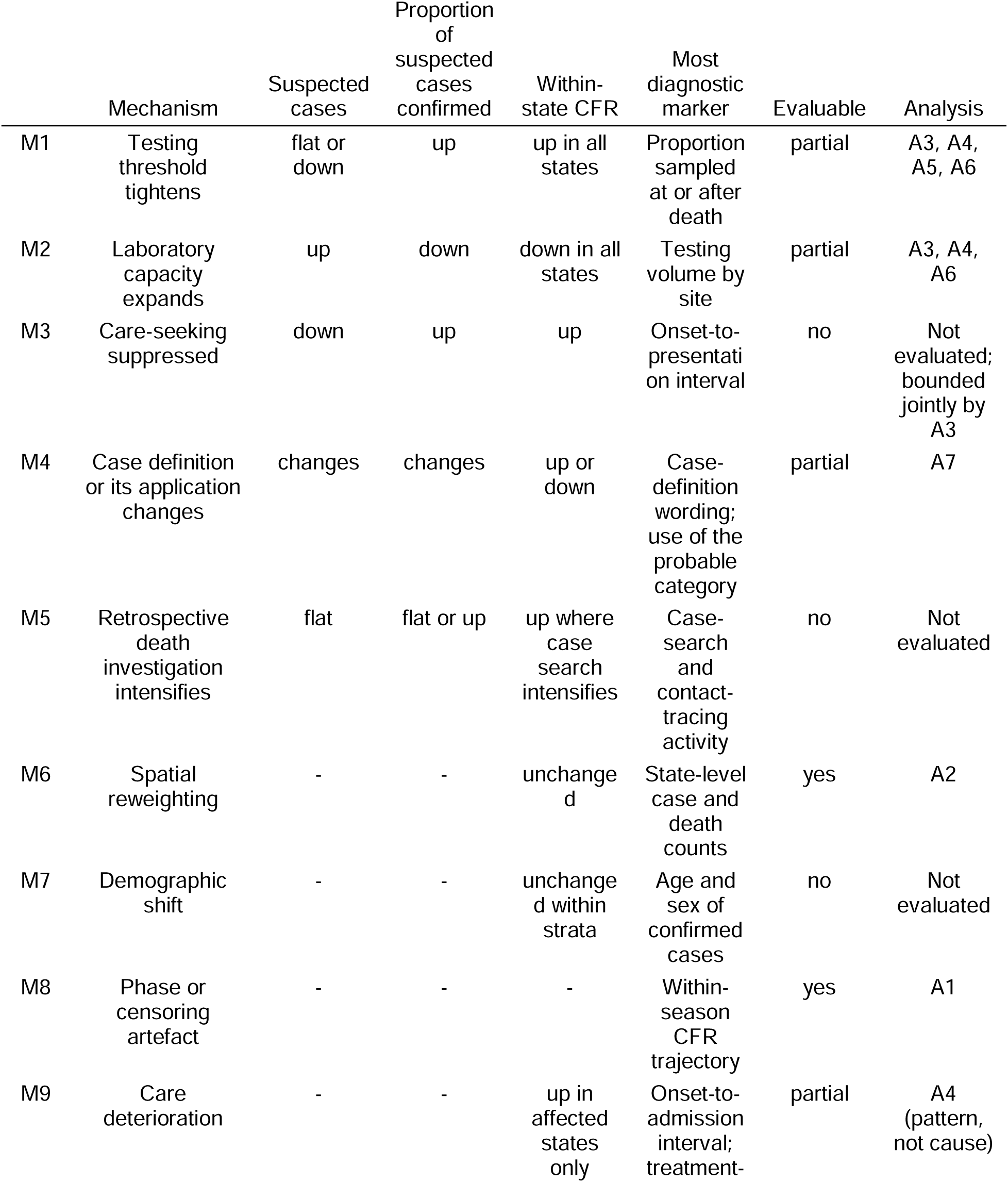
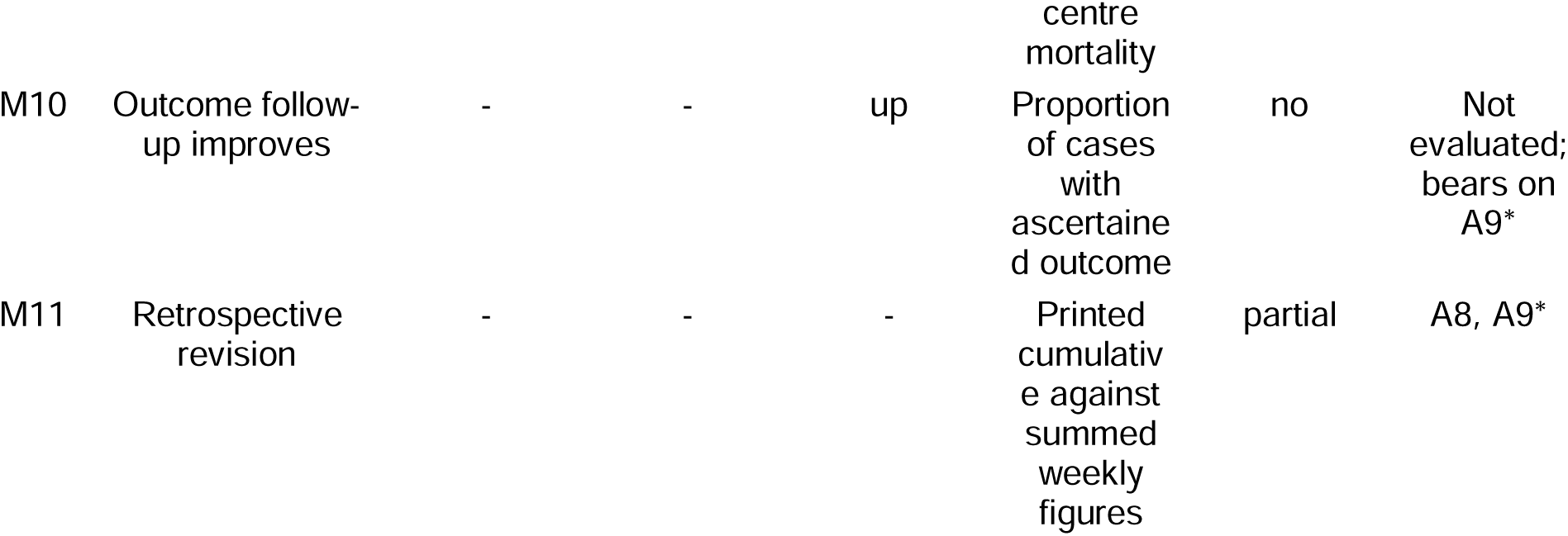
Mechanisms that can move a reported CFR, the signature each would leave, and whether published NCDC reports allow it to be evaluated. Analyses A1-A8 are numbered as in the pre-specified plan. *A9 (printed cumulative against summed weekly figures) was added after the plan was fixed; evaluability is shown as pre-specified.

### Analyses

Each analysis evaluates one or more rows of Table 1 and has one main estimand (Table 2). The conclusions rest on three primary analyses. Two pre-specified secondary analyses, a hierarchical model of common against localised change and a test of detection triggered by death, are reported in the appendix.

**Table 2:** Main estimands of the primary and secondary analyses, and the main results of the analyses added after the plan was fixed. Conclusions rest on the primary analyses; intervals are 95% CIs.

| Role | Analysis | Estimand | Estimate | Interval |
| --- | --- | --- | --- | --- |
| Primary | A1 Phase-matched comparison | 2026 minus 2025 cumulative CFR at the 50% phase point | +0.3 percentage points | -5.0 to 7.6 |
| Primary | A2 Decomposition | Between-state component of the 2025-2026 change | +0.5 percentage points | -2.0 to 2.9 |
| Primary | A2 Decomposition | Within-state component of the 2025-2026 change | -1.1 percentage points | -4.7 to 3.0 |
| Primary | A3 Tipping point | Missed survivors at week 23 as a share of suspected cases not confirmed | 6% | 2% to 11% |
| Secondary | A4 Hierarchical model | P(year SD > state-within-year SD) | 0.14 | - |
| Secondary | A5 Isolated detection | Observed / expected share with a death | 1.18 | 0.90 to 1.30 |
| Added after the plan was fixed | A9 Printed against summed weekly figures | Deaths added outside the weekly figures, 2026 to week 36 | 70 | - |
| Added after the plan was fixed | Addendum 3: phase-matched comparison, published series | 2026 minus 2025 cumulative CFR at the 50% phase point | +5.4 percentage points | - |

#### Phase-matched comparison

Deaths lag cases and seasons rise at different speeds, so a cumulative CFR read on the same calendar week can compare years at different points in their seasons [2]. I compared cumulative CFR at the week each year reached 50% (primary) and 25% of the confirmed cases it had by week 36, the last week reported in 2026. The fixed week 23 comparison is shown alongside.

#### Decomposition

A national CFR is a case-weighted average of state CFRs, so it can rise because more cases occur in states where the CFR has long been higher, without the risk of death changing anywhere. I split each year-on-year change from 2018 into a within-state and a between-state component [18], with every year cut to week 36. For each pair of years a state was its own stratum if it had at least 30 confirmed cases in both, a threshold set before analysis to avoid unstable CFRs from small counts, and on cases rather than deaths so that strata were not chosen on the outcome; the rest were pooled. The within-state component still combines changes in the risk of death with changes in who is tested.

#### Tipping point

I computed how many survivors would have to be missing (people with symptomatic Lassa fever who survived without being confirmed) from the 2026 confirmed cases at week 23 for its CFR to equal that of 2025, using the published figures the commentary quoted, and expressed the number as a share of the 2026 suspected cases that were not confirmed.

#### Post-protocol exploratory analyses

After the initial results, three addenda were added. First, a comparison of the cumulative figures printed in each report with the summed weekly figures, by year, state and week. Second, a repeat of the annual and phase-matched comparisons on the printed series, as point estimates. Finally, a check of rebuilt and missing weeks against an independent extraction of NCDC’s Weekly Epidemiological Report was performed [19].

#### Uncertainty and software

Intervals for the primary analyses come from a bootstrap that resamples state-weeks within each year, with 1,000 replicates, and are reported as 95% confidence intervals (CIs); the model reports 95% credible intervals (CrIs). Analyses used R 4.6 with brms and cmdstanr (appendix); all code and the analysis plan are available (see Data availability statement).

#### Use of AI tools

AI tools (Claude Opus 5.5, Anthropic) were used to develop, test and comment the analysis code, to compare the completed extraction with the machine-readable parts of each report, and to revise text. Every value was reviewed, and I checked all code, outputs and text.

#### Sex and gender

The situation reports give no breakdown of cases or deaths by sex or age, so neither could be analysed; the Discussion considers what this omission means for interpreting the CFR.

#### Patient and public involvement

Patients and the public were not involved in the design, conduct, reporting or dissemination of this research, which used published aggregate surveillance reports only.

## Results

From 2018 to week 36 of 2026, the reports recorded 8,811 confirmed cases and 1,473 deaths among them, in every state and the Federal Capital Territory. No report exists for 8 weeks, and five weeks of 2022 survive only as a single total (appendix).

### Evaluability of candidate mechanisms

Of the 11 mechanisms in Table 1, 2 can be evaluated using published situation reports, 5 in part, and 4 not at all. The unreported variables include testing volume, clinical intervals (symptom onset to presentation, and diagnosis to death), the proportion of cases sampled post-mortem, outcome-ascertainment completeness, admitting healthcare facility tier, and the age and sex of patients. Without these, the mechanisms that raise the CFR within states cannot be distinguished: tighter clinical testing thresholds (M1), suppressed health-seeking behaviour (M3), more intensive post-mortem investigation (M5), deteriorating clinical care (M9) and more complete mortality assessment (M10) generate similar signatures in aggregated data.

### Discrepancy between cumulative and weekly death counts

Each report gives the figures for its week and a cumulative total for the year, and the two disagree on deaths. In every year permitting like-for-like comparison, cumulative confirmed cases closely matched the sum of the weekly figures, but cumulative deaths exceeded the summed weekly deaths, by 94 in 2020, 45 in 2022, 17 in 2023, 5 in 2024, 4 in 2025, 70 in 2026 (to week 36) (Figure 1). In 2026 47 of the 70 added deaths were in Ondo, and nationally they were added in batches of five or more deaths at weeks 6, 7, 10, 17, 19, 33 rather than appearing in any week’s figures. The published CFR is calculated from these cumulative totals, as in the national surveillance guidelines and NCDC’s CFR target [14,20].

**Figure 1:**
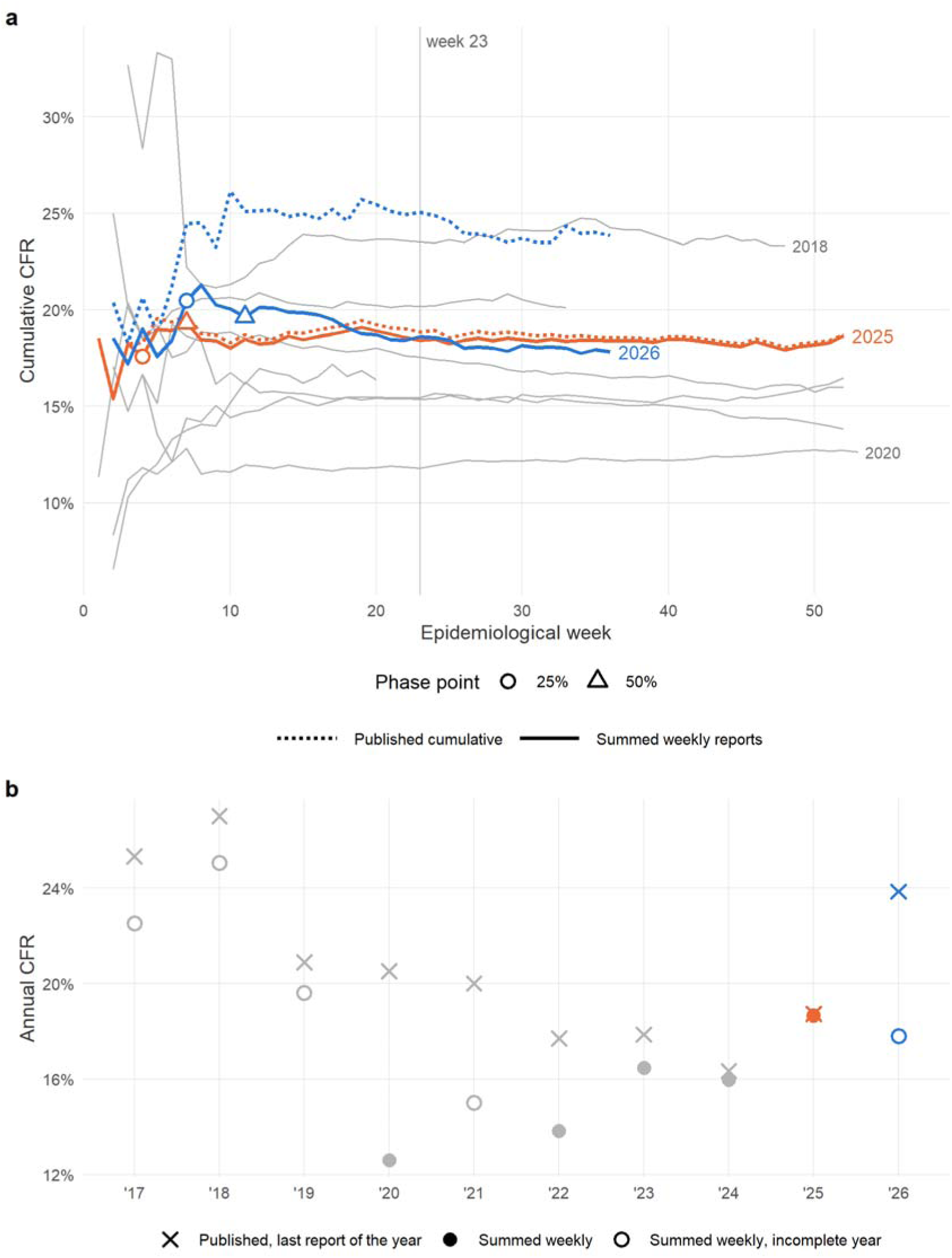
Reported case fatality in two series: the running sum of the weekly reports (solid lines, filled and hollow points), used by the pre-specified analyses, and the cumulative figures printed in each report (dashed lines, crosses), which also hold deaths added after the week they occurred (Addendum 3, added after the analysis plan was fixed). (a) Cumulative CFR through each year from the week the year reached 50 confirmed cases, with 2025 and 2026 highlighted and the 25th and 50th percentile phase points (summed weekly series); the vertical line is week 23. Grey lines are earlier years; a line stops at the first week with no report, after which the running total is unknown. (b) Annual CFR, 2017-2026. Summed weekly: deaths and confirmed cases from weeks in which both are reported, hollow where the year has missing weeks or is incomplete. Published: each year’s final printed cumulative figure, for 2026 the week 36 report; the 2017 figure counts from December 2016.

Between 2020 and 2025 the published annual CFR ranged from 16.3% to 20.5%, whereas the CFR derived from weekly reports ranged from 12.6% to 18.7%. By week 36 of 2026, the published cumulative CFR was 23.8% compared with 17.8% from the weekly incident series.

### Phase-matched comparisons

On the weekly series, the week 23 comparison quoted in the commentary gave a CFR of 18.4% in 2025 and 18.6% in 2026, a difference of +0.2 percentage points (95% CI - 4.3 to 5.1). The two years reached half of their confirmed cases to week 36 at weeks 7 and 11; the difference at this point was +0.3 percentage points (95% CI -5.0 to 7.6), and at the 25th percentile point +2.9 percentage points (95% CI -7.0 to 11.8). By contrast, the same comparisons using the published cumulative series gave +6.2 percentage points, +5.4 percentage points and +6.2 percentage points (point estimates). Phase matching made little difference: whether 2025 and 2026 differ depends on which count of deaths is used, not on when in the year the two are compared.

### Spatial decomposition of case fatality

On the weekly series to week 36, the 2025-2026 change in national CFR was -0.6 percentage points (95% CI -4.7 to 3.7), an interval that included zero. The within-state component was -1.1 percentage points (95% CI -4.7 to 3.0) and the between-state component +0.5 percentage points (95% CI -2.0 to 2.9) (Figure 2). 4 high-burden states met the threshold to stand alone in this paired comparison (Bauchi, Edo, Ondo, Taraba). With every state separate (Figure 2, panel b), the largest contributions came from Bauchi, Edo and the growth of Benue’s cases; each rests on few cases, and their intervals all included zero (appendix). Across all 8 transitions, 3 within-state components had intervals excluding zero (2019-2020, 2020-2021, 2024-2025); with 8 comparisons and intervals that are probably too narrow, these are not individually conclusive.

**Figure 2:**
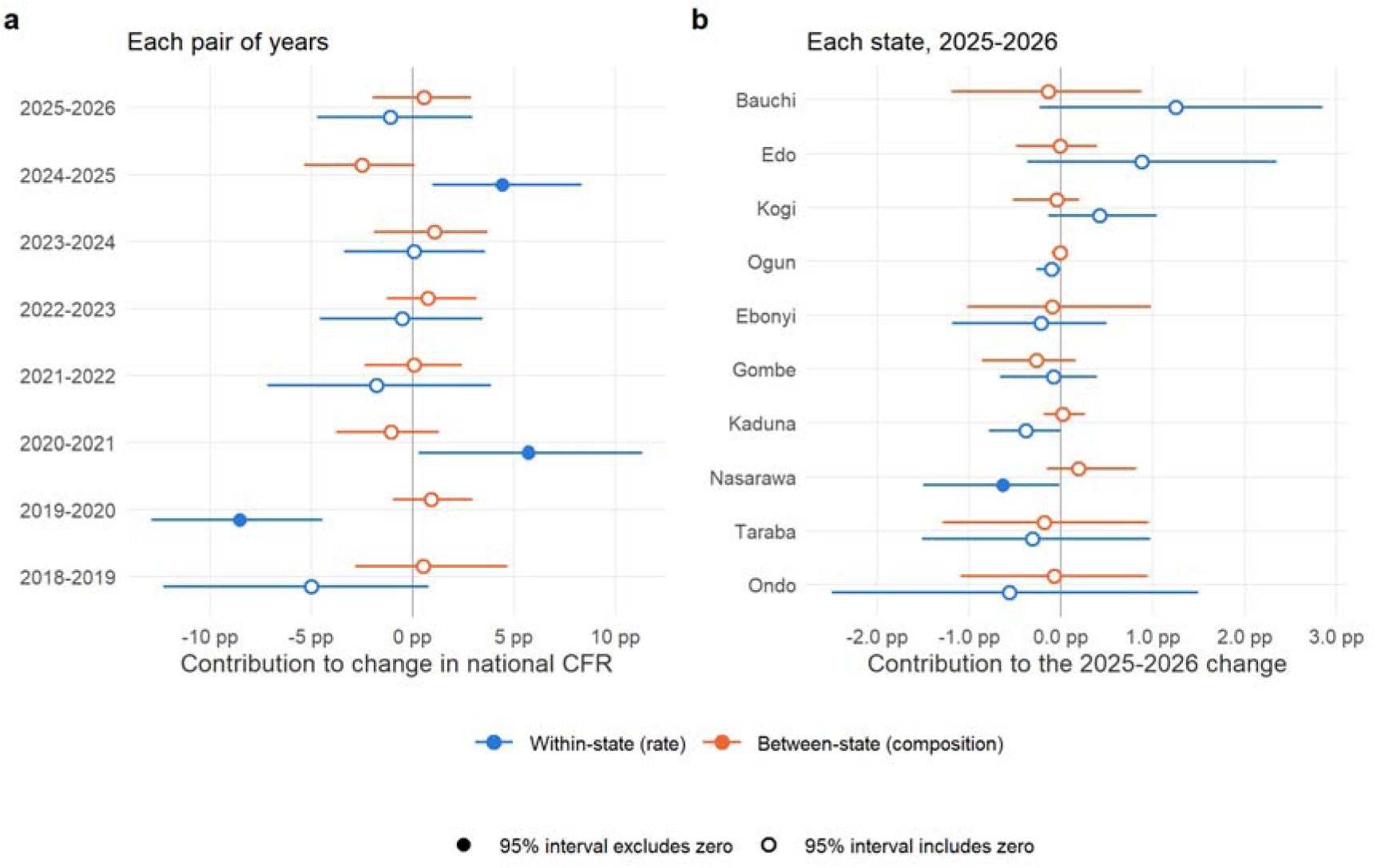
Decomposition of year-on-year change in national CFR (summed weekly reports, every year cut to week 36). (a) Within-state and between-state components for every pair of consecutive years. (b) Each state’s contribution to the 2025-2026 change, every state separate: a secondary output, imprecise because each state contributes few cases. The two components add up to the observed change and need not share a direction. Bars are 95% bootstrap intervals; points are hollow where the interval includes zero. The panels have different scales.

### Tipping point for ascertainment

To reduce the published 2026 cumulative CFR at week 23 from 25.0% to the 2025 level of 18.9% would require 279 survivors to have been missed (95% CI 91 to 505), 33% of the confirmed cases and 6.1% of the suspected cases that were not confirmed (95% CI 2.0% to 11.0%) (Figure 3). On the weekly series it would take 9 (95% CI -134 to 182). On the published figures this is about 12 survivors a week; on the weekly figures it is under one a week (0.4), with an interval that included zero.

**Figure 3:**
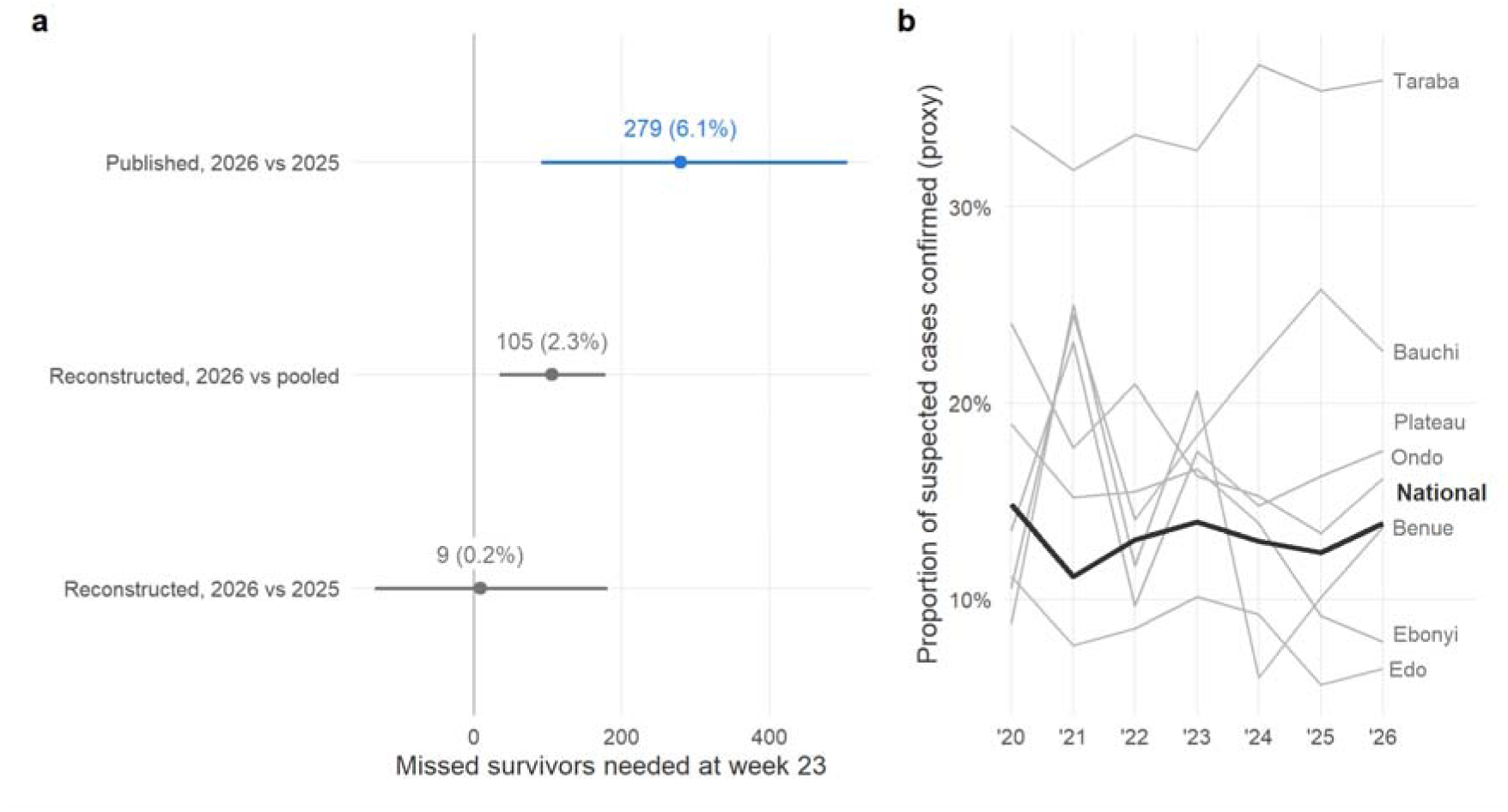
Ascertainment. (a) The number of survivors who would have to be missing from the 2026 confirmed cases at week 23 for the 2026 CFR to equal the reference CFR, with 95% bootstrap intervals; labels give the number and, in brackets, its share of the 2026 suspected cases that were not confirmed by week 23, the main estimand. Published: the cumulative figures quoted by the commentary (primary). Reconstructed: the summed weekly reports. Pooled: all earlier years from 2018 whose week 23 total is exactly known. (b) Proportion of suspected cases confirmed (a proxy that includes the decision to test), nationally (black) and for the states that stand alone in any decomposition pair (grey), from 2020, when suspected cases are given by state.

### Case definitions and the proportion of suspected cases confirmed

The confirmed-case definition remained identical throughout the study period, as did clinical criteria for suspected cases. A suspected case who died before a specimen was taken was classified as probable throughout (from week 25 of 2019, also one who absconded), so deaths before sampling are excluded from both counts of the reported CFR. The probable category was used in 7 of 9 years, by at most 5 states in any single year. Nationally, the proportion of suspected cases confirmed was 12.4% in 2025 and 13.9% in 2026, and changed little from 2020 apart from a fall in 2021, but it differed widely and persistently between states, from 5.6% to 11.2% in Edo, falling over the period, to 31.9% to 37.2% in Taraba (Figure 3, panel b).

### Secondary analyses

The hierarchical model yielded inconclusive evidence regarding common versus localised inter-annual variance: the posterior probability that the year standard deviation exceeded the state-within-year standard deviation was 0.14 under a binomial likelihood, in which the state-within-year term also absorbs overdispersion, and 0.71 with a beta-binomial, which carries it in a separate term. Isolated detections were followed by a death 1.18 times as often as expected under severity-blind detection (95% CI 0.90 to 1.30) (appendix).

## Discussion

The rise in Nigeria’s reported Lassa fever CFR in 2026 reflects how cumulative totals are updated over time, not an increase in deaths in the weekly incident reports. Calculated from the weekly reports, the 2026 CFR was indistinguishable from 2025, at week 23 and at every phase-matched point. The headline increase came instead from 70 deaths, 47 of them in Ondo State, added to the cumulative totals by week 36 without appearing in any week’s count. Far fewer confirmed cases than deaths were added in the same way (25 against 70), so most of these deaths were probably among people already counted as confirmed cases in earlier weeks. This points to delayed outcome reporting and routine reconciliation of records (M10, M11 in Table 1), not to deaths found later by case search (M5). Seasonal timing does not explain the difference, which persisted at every phase-matched point, and the movement of cases between states did not change the weekly figures. About 12 missed survivors a week (6% of suspected cases not confirmed by week 23) could produce a rise of the same size, but the published reports do not contain the testing denominators needed to tell whether ascertainment narrowed.

These findings do not show that clinical care in 2026 matched 2025, only that published surveillance figures cannot show that it worsened. Attributing surveillance artefacts to clinical failure has the potential to misdirect resources toward referral pathways when access to testing may be the primary constraint; conversely, dismissing genuine clinical decline leaves avoidable mortality unaddressed.

This analysis was possible because NCDC publishes, every week, state-level counts of suspected, confirmed and probable cases and of deaths, with cumulative totals, and keeps every report online. Box 1 translates these mechanisms into a framework to evaluate changes in reported CFR, each paired with the data that would answer it. The mechanisms the published reports could evaluate fully or in part come down to four operational questions: whether years are compared at the same point in the season (M8), whether a change exceeds chance variation, whether cases moved between states (M6), and how many deaths were added after the week of report (M10, M11; median 31 a year, range 4 to 94). However, public reports could not answer whether testing changed, whether fatal cases were identified preferentially post-mortem, whether clinical presentation was delayed, or whether the age profile shifted, an important consideration because CFR was higher in older patients in each state whose records have been analysed [13,21,22]. Because surveillance CFR conflates these variables with quality of care, progress toward the national strategic target of a CFR below 10% [14] cannot be judged from the figure alone.

For Lassa fever, the suspected-case definition provides limited screening specificity in endemic regions: in a West African multicentre cohort, only 39 of 7,321 illnesses that met these criteria were confirmed Lassa fever [7], and the definition’s exposure criterion, contact with rodents or their excreta, excludes few people in endemic areas, where 94% of rural households in Nigeria report rodents entering the home [23]. Suspected-case identification relies on clinical discretion, so the proportion of suspected cases confirmed combines how often a suspected case is tested with how often a test is positive; it is sometimes read as a laboratory confirmation rate [24]. Where state records hold test results, true test positivity can be followed: in Ebonyi it fell from above 40% in the peak months of 2018-2019 to 10-20% in 2020-2023, while the state’s CFR rose from 16% to 57% [21], a combination the published figures alone cannot explain.

Health-system strain biases reported CFR in opposing directions. Patients dying prior to biological sampling are classified as probable cases and removed from both numerator and denominator, depressing the reported ratio; conversely, rationing diagnostics to severe inpatients or testing post-mortem inflates it. Because the probable classification was rarely used, deaths before sampling may be going unrecorded rather than not occurring.

The two counts of deaths reflect how case-based surveillance works, not errors in reporting. Cases are recorded individually in SORMAS, the electronic surveillance platform introduced in 2018 [21,22], and the national surveillance guidelines direct that late reports be entered in their own week, with totals updated [20]. As outcomes are completed and records reconciled, deaths can reach the cumulative totals after the week in which the case was reported (K Jan, Nigeria Centre for Disease Control and Prevention, personal communication), so the cumulative series is the more complete count of deaths. But the number added outside the weekly figures varies widely between years and states. Few deaths were added in 2024 and 2025 and many in 2026, mostly in batches from one state; this could reflect slower notification of deaths or more thorough reconciliation of records, and the reports do not show which. Even the year-end CFR is therefore not comparable from year to year unless late additions are published by week of confirmation, and analyses of these reports should state which series they use. The guidelines also record a case’s outcome without an observation window [20], so a death from a later complication may or may not be counted, as debated for COVID-19 as deaths from against deaths with the disease [25].

Others have called for care-pathway intervals to be reported [16]. They would show whether patients reach care later only if defined consistently, because symptom onset is recalled, sometimes by patients in extremis, and fever may go unrecorded [7]. Separating a change in care from a change in ascertainment also requires published testing volumes, the proportion of cases first sampled at or after death, and the share of confirmed cases with a known outcome.

The same figures are used to plan trials. Lassa fever vaccine candidates are now entering phase 2 trials [26], and the WHO target product profile sets efficacy against infection or disease as the measure while noting that incidence estimates are biased by limited testing [15]. An efficacy trial needs an expected number of confirmed cases at its sites and, for a severe-disease or mortality endpoint, an expected CFR; planning models calibrated to surveillance counts inherit whatever moves those counts [27].

Confirmed cases in Edo fell from 351 in 2023 to 185 in 2025 while suspected cases held steady, so a site ranked on confirmed cases may reflect where testing is done rather than where infection occurs. Health-seeking can shift in the same way: presentations at Kenema in Sierra Leone fell after the 2013-16 Ebola epidemic through changes in health-seeking rather than in transmission [28]. A reported CFR that rises because deaths are reconciled late, or because testing narrows to the sickest patients, would overstate the severe outcomes a trial can expect, and one that falls as testing widens would understate them. Therapeutic trials restricted to mild or moderate disease [29] enrol from the confirmed cases surveillance finds, so their expected mortality cannot be read from a surveillance CFR either. Linking trial site selection to surveillance and ecological data, as proposed in a One Health trial design [30], depends on separating testing denominators and record reconciliation from incidence and severity; the reporting set in Box 1 would let trialists and funders do so. Simulating trial accrual under an explicit model of surveillance would show how much this matters.

This study has limitations. All inference is ecological. Numerator ascertainment could be tracked only through the difference between the two death series, and the spatial decomposition allocates changes arithmetically without establishing clinical causality. The bootstrap ignores correlation between a state’s consecutive weeks, so its intervals are probably too narrow, and comparing consecutive years can miss a slow drift in ascertainment. National counts agreed closely with an independent extraction [19], but the reports are a snapshot of a case-based system whose records are updated after publication: state and treatment-centre records give different counts for the same years [13,21], and an analysis of the line lists themselves would show how the published figures relate to the records they summarise.

The difficulty is not specific to Lassa fever. In other zoonoses with recurrent spillover the reported CFR also depends on which cases are confirmed, and the same questions apply: who is tested, which deaths are counted, and whether sampling depends on the outcome. In Afghanistan the CFR of Crimean-Congo haemorrhagic fever was 43% among confirmed cases and 14% among suspected cases in 2016-18 [31]; the choice of denominator alone tripled the figure, so the confirmed CFR moves with the share of suspected cases that are tested. In Bangladesh a quarter of Nipah virus cases are probable, cases that died before a specimen could be taken [32]; whether such deaths enter the CFR depends on how cases are classified, and a change in how quickly fatal cases are sampled would shift it. In Madagascar’s 2017 epidemic plague was confirmed more often among deaths, because specimens taken after death gave a higher yield [33], so wider post-mortem sampling would raise the confirmed CFR without any change in risk. Crimean-Congo haemorrhagic fever and Nipah virus infection, like Lassa fever, are WHO priority diseases for countermeasure development [1], so their reported CFRs carry the same weight in setting targets and planning trials. Publishing the quantities in Box 1 would let a change in reported CFR be read as a change in outcome, or ruled out as one.

## Conclusion

Nigeria’s reported Lassa fever CFR is used to track a national target and to plan trials, but in 2026 its rise came from deaths added to cumulative totals, not from the weekly figures, and the published data cannot show whether care changed. The data that would show it are already collected in case-based surveillance. Publishing them as a minimum reporting set would let CFR targets be tracked, give trialists and funders baselines that separate surveillance from severity, and do the same for other endemic zoonoses.

## Data Availability

The situation reports are public on the NCDC website. The extracted data, analysis code, pre-specified analysis plan and its dated addenda are available at https://github.com/DidDrog11/lassa-cfr and archived at Zenodo, where doi:10.5281/zenodo.23135367 resolves to the latest version.

https://doi.org/10.5281/zenodo.23135367

## Contributors

DS was responsible for conceptualisation, data curation, formal analysis, investigation, methodology, project administration, software, validation, visualisation, and writing (original draft, review and editing). DS accessed and verified the underlying data, had final responsibility for the decision to submit for publication, and is the guarantor.

## Patient consent for publication

Not applicable.

## Ethics approval

Not applicable. The study used only publicly available, aggregated surveillance reports and involved no human participants.

## Data availability statement

The situation reports are public on the NCDC website. No individual participant data were collected; the study uses aggregate counts only. The extracted data (state-by-week counts and the printed cumulative figures, with the extraction checks), the analysis code, the pre-specified analysis plan and its dated addenda are available now, without restriction, under a CC0 licence at https://github.com/DidDrog11/lassa-cfr, and archived at Zenodo, where doi:10.5281/zenodo.23135367 resolves to the latest version. The repository README describes each data file.

## Acknowledgements

I thank Dr Kamji Jan (Nigeria Centre for Disease Control and Prevention) for explaining how cases and deaths are recorded and reported in NCDC’s Lassa fever situation reports. The views expressed are the author’s own and do not necessarily reflect those of NCDC.

## Declaration of generative AI use

During this work I used Claude Opus 5.5 (Anthropic) to develop, test and comment the analysis code, to compare the completed extraction with the machine-readable parts of each report, and to revise text. Every value was manually extracted from the reports, and corrections suggested by AI tools were individually confirmed. I reviewed and edited all code, outputs and text produced with the tool, and take full responsibility for the content of the publication.

## Box 1: A proposed minimum reporting set for case fatality in endemic zoonotic surveillance

A reported case fatality ratio (CFR) is a ratio of two surveillance counts, not an intrinsic biological parameter. In endemic zoonoses with seasonal spillover, comparisons between years conflate clinical outcome with shifting access to care, testing thresholds and reporting practices. Each question below is paired with the data that would answer it. In Nigeria most of these data are already recorded for each case in SORMAS, the case-based surveillance platform, which holds sample dates and results, outcome and date of death, place of detection and age group,^21^ so they could be published as weekly counts by state alongside the existing situation report tables.

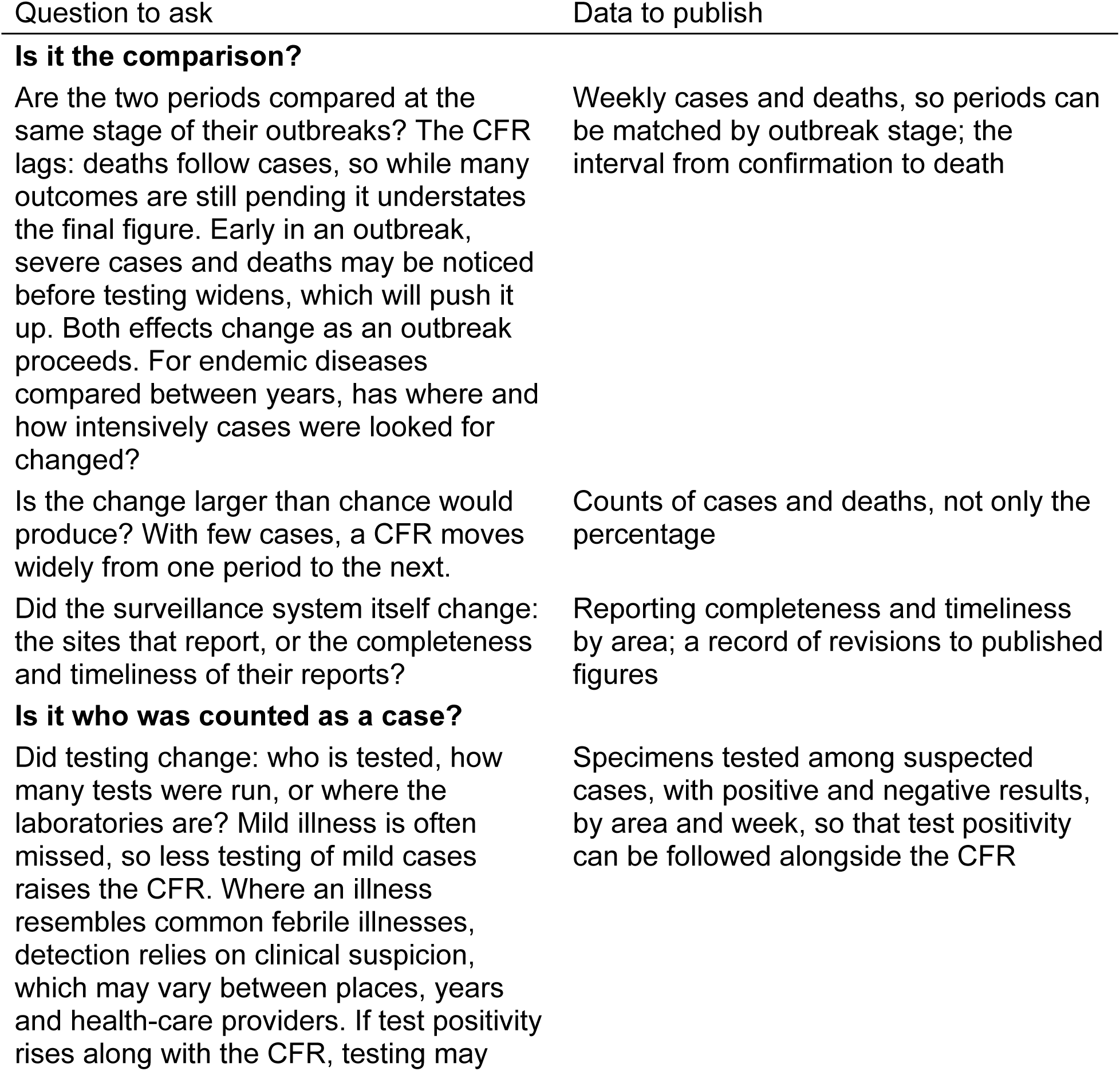
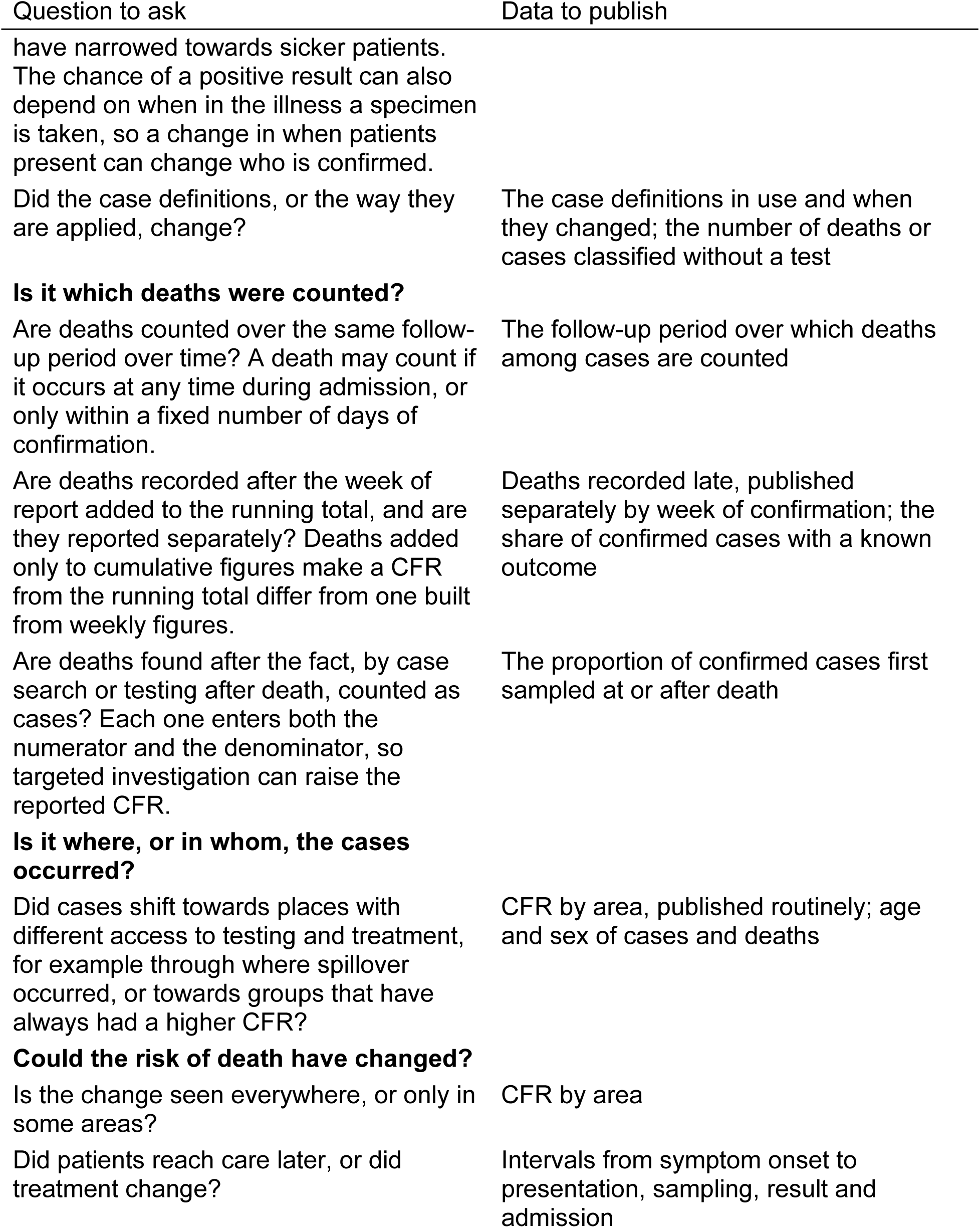

## References

1 World Health Organization. 2017 annual review of diseases prioritized under the Research and Development Blueprint: Informal consultation, 24-25 January 2017, Geneva, Switzerland. Meeting report. Geneva: World Health Organization 2017.

2 Lipsitch M, Donnelly CA, Fraser C, et al. Potential biases in estimating absolute and relative case-fatality risks during outbreaks. PLoS Neglected Tropical Diseases. 2015;9:e0003846. doi: 10.1371/journal.pntd.0003846

3 Reich NG, Lessler J, Cummings DAT, et al. Estimating absolute and relative case fatality ratios from infectious disease surveillance data. Biometrics. 2012;68:598– 606. doi: 10.1111/j.1541-0420.2011.01709.x

4 Verity R, Okell LC, Dorigatti I, et al. Estimates of the severity of coronavirus disease 2019: A model-based analysis. The Lancet Infectious Diseases. 2020;20:669–77. doi: 10.1016/S1473-3099(20)30243-7

5 Lloyd-Smith JO, George D, Pepin KM, et al. Epidemic dynamics at the human-animal interface. Science. 2009;326:1362–7. doi: 10.1126/science.1177345

6 Kock R, Caceres-Escobar H. Situation analysis on the roles and risks of wildlife in the emergence of human infectious diseases. Gland, Switzerland: IUCN 2022.

7 Issifou AMD, Deha ARE, Yadouleton A, et al. Incidence of symptomatic Lassa virus infection in West African countries (Enable 1.0 Lassa Research Study): A prospective, multisite, cohort study. The Lancet Global Health. 2026;14:103945. doi: 10.1016/S2214-109X(26)00090-2

8 Hegde ST, Salje H, Sazzad HMS, et al. Using healthcare-seeking behaviour to estimate the number of nipah outbreaks missed by hospital-based surveillance in bangladesh. International Journal of Epidemiology. 2019;48:1219–27. doi: 10.1093/ije/dyz057

9 Dalhat MM, Olayinka A, Meremikwu MM, et al. Epidemiological trends of Lassa fever in Nigeria, 2018–2021. PLOS ONE. 2022;17:e0279467. doi: 10.1371/journal.pone.0279467

10 McCormick JB, Webb PA, Krebs JW, et al. A prospective study of the epidemiology and ecology of Lassa fever. Journal of Infectious Diseases. 1987;155:437–44. doi: 10.1093/infdis/155.3.437

11 Simons D. Lassa fever cases suffer from severe underreporting based on reported fatalities. International Health. 2022;15:608–10. doi: 10.1093/inthealth/ihac076

12 Doohan P, Jorgensen D, Naidoo TM, et al. Lassa fever outbreaks, mathematical models, and disease parameters: A systematic review and meta-analysis. The Lancet Global Health. 2024;12:e1962–72. doi: 10.1016/S2214-109X(24)00379-6

13 Cadmus S, Awosanya E, Iziomo PM, et al. Deciphering the dynamics of Lassa fever outbreak in Edo State, Nigeria: A 3-year study of the epidemiologic patterns, severity profiles, and clinical management. IJID One Health. 2025;7:100059. doi: 10.1016/j.ijidoh.2025.100059

14 Nigeria Centre for Disease Control and Prevention. Advocacy toolkit: Influencing key decision-makers to create an enabling environment for Lassa fever preparedness and response in Nigeria. Abuja: Nigeria Centre for Disease Control; Prevention 2025.

15 World Health Organization. WHO target product profile for Lassa virus vaccine. Geneva: World Health Organization 2017.

16 Abdulrahim A, Gulumbe BH, Adepoju VA. Increasing mortality during Nigeria’s 2026 Lassa fever outbreak calls for earlier diagnosis and treatment. The Lancet Regional Health – Africa. 2026;100135. doi: 10.1016/j.lanafr.2026.100135

17 Simons D. Map liberator. 2026.

18 Kitagawa EM. Components of a difference between two rates. Journal of the American Statistical Association. 1955;50:1168–94. doi: 10.1080/01621459.1955.10501299

19 Niyi-Oriolowo E. NCDC lassa fever weekly timeseries dataset (nigeria, 2020-2025). 2025.

20 Federal Ministry of Health, Nigeria Centre for Disease Control. National technical guidelines for integrated disease surveillance and response. 3rd edn. Abuja: Federal Ministry of Health 2019.

21 Iziomo PM, Awosanya E, Taiwo OJ, et al. Temporal trends and epidemiological patterns of Lassa fever outbreaks in Ebonyi State, Nigeria: A retrospective study (2018-2023). PLOS Global Public Health. 2026;6:e0006871. doi: 10.1371/journal.pgph.0006871

22 Cadmus S, Akinseye V, Cadmus E, et al. Drivers of Lassa fever in an endemic area of southwestern Nigeria (2017-2021): An epidemiological study. PLoS One. 2025;20:e0327143. doi: 10.1371/journal.pone.0327143

23 Simons D, Harden C, Imirzian N, et al. Local heterogeneity in Lassa fever serology in rural Nigeria: Implications for vaccine trial site selection. PLoS Neglected Tropical Diseases. 2026;20:e0014379. doi: 10.1371/journal.pntd.0014379

24 Tadema AJ, Nwosu FM, Abimbola OA. Temporal dynamics, climate associations, and surveillance performance of lassa fever in nigeria, 2020–2025: A national longitudinal analysis. Osong Public Health and Research Perspectives. 2026;17:310–21. doi: 10.24171/j.phrp.2026.0226

25 Buschner A, Katz K, Beyerlein A. Comparison of fatalities due to COVID-19 and other nonexternal causes during the first five pandemic waves. Bundesgesundheitsblatt - Gesundheitsforschung - Gesundheitsschutz. 2024;67:939–46. doi: 10.1007/s00103-024-03914-5

26 Bardach A, Berrueta M, Ciapponi A, et al. Efficacy, safety, and immunogenicity of Lassa fever vaccines: A living systematic review and landscape analysis of vaccine candidates. PLoS One. 2025;20:e0338128. doi: 10.1371/journal.pone.0338128

27 Moore SM, Rapheal E, Mendoza Guerrero S, et al. Estimation of Lassa fever incidence rates in West Africa: Development of a modeling framework to inform vaccine trial design. PLoS Neglected Tropical Diseases. 2025;19:e0012751. doi: 10.1371/journal.pntd.0012751

28 Koch MR, Kanneh L, Wise PH, et al. Health seeking behavior after the 2013-16 Ebola epidemic: Lassa fever as a metric of persistent changes in Kenema District, Sierra Leone. PLoS Neglected Tropical Diseases. 2021;15:e0009576. doi: 10.1371/journal.pntd.0009576

29 Erameh C, Okwaraeke K, Kleist C, et al. Favipiravir for Lassa fever: An open-label, randomized controlled phase 2 trial. Nature Medicine. Published Online First: 2026. doi: 10.1038/s41591-026-04402-w

30 Simons D, Watson-Jones D, Manno D, et al. A One Health trial design to accelerate Lassa fever vaccines. Nature Medicine. 2026;32:25–7. doi: 10.1038/s41591-025-04018-6

31 Sahak MN, Arifi F, Saeedzai SA. Descriptive epidemiology of Crimean-Congo hemorrhagic fever (CCHF) in Afghanistan: Reported cases to National Surveillance System, 2016–2018. International Journal of Infectious Diseases. 2019;88:135–40. doi: 10.1016/j.ijid.2019.08.016

32 Satter SM, Aquib WR, Sultana S, et al. Tackling a global epidemic threat: Nipah surveillance in Bangladesh, 2006–2021. PLOS Neglected Tropical Diseases. 2023;17:e0011617. doi: 10.1371/journal.pntd.0011617

33 Randremanana R, Andrianaivoarimanana V, Nikolay B, et al. Epidemiological characteristics of an urban plague epidemic in Madagascar, August–November, 2017: An outbreak report. The Lancet Infectious Diseases. 2019;19:537–45. doi: 10.1016/S1473-3099(18)30730-8

