## Appendix for "Reported case fatality as a target and trial metric in endemic zoonoses: a framework and ecological analysis of Lassa fever surveillance in Nigeria, 2017–2026"

David Simons

#### Supplementary methods

*Reporting.* The study is reported following the RECORD statement for observational studies using routinely collected health data[1]; the completed checklist is in Table S2.

*Literature search.* PubMed and Europe PMC were searched on 13 October 2026 for the Research in context panel, in titles and abstracts, with no date or language restriction. Four groups of terms were combined with AND: case fatality (`"case fatality"`, `"case-fatality"`, `"fatality ratio*"`, `"fatality rate*"`); bias or ascertainment (`ascertain*`, `under-ascertain*`, `bias*`, `artefact*`, `artifact*`, `underreport*`, `under-report*`, `censor*`, `delay*`); change or interpretation (`trend*`, `temporal`, `seasonal*`, `secular`, `decompos*`, `interpret*`, `change*`, `comparison*`); and surveillance (`surveillance`, `reported`, `reporting`). PubMed returned 266 records and Europe PMC 288, of which 59 (53 preprints) were not in the PubMed set. Records were included if they formally separated a change in a reported CFR over time from mechanisms other than outcome (by decomposition, standardisation, or adjustment for testing, ascertainment, reporting delay or death reporting) and discussed what the change could show about outcome, or if they evaluated whether routine surveillance data could support reading such a change as a change in outcome. Reporting a CFR trend, or naming ascertainment as a caveat, was not enough. The author screened the titles and abstracts of the first 30 records (about 10%); the remaining 295 were screened with AI assistance against the same criteria. The author reviewed every record marked for inclusion or full-text review, and a random 25% of the records excluded with AI assistance (69 of 274). The author confirmed all 69 sampled exclusions. Of 325 records, 10 met the criteria on title and abstract and 9 needed the full text; all 9 full texts were read, and 2 met the criteria. Of the 12 studies included, 11 concerned COVID-19 and one meningococcal disease, both spread in chains of human transmission; none concerned a zoonosis with recurrent spillover.

*Extraction and data handling.* The completed extraction was checked against the machine-readable parts of each report (national totals on page one from 2020, and the state-by-state enumeration in the highlights text for 2017-2019), and every discrepancy was resolved against the report. Across 2018-2026 the checks found 80 of 18,152 extracted values (0.4%) entered incorrectly, between 0.1% and 1.9% a year (Table S1), all corrected before the final run; in a further 15 weeks the national totals on page one disagreed with the report's own state table, and the state table was kept. 2017 was not checked. The checks find omitted and mistyped values, but not errors that cancel out or a value entered under the wrong state. From week 7 of 2020 each report carries a state table; for 2017-2019 state figures come from the highlights text, which gives confirmed cases and deaths by state but not suspected or probable cases in 2018-2019. Only states named in a report were extracted; a state absent from a report that listed a variable by state was taken as zero, and one absent from a report without a state breakdown as missing. Weeks with no report, whether never published or missing from the archive, were left missing. Cases reported late in 2017-2019 were added to the week of the report that carried them, and the 2017 list of suspected cases by state was taken as complete. Deaths reported only as a national total, in seven reports in 2018 and 2020, were allocated to states with confirmed cases that week in proportion to their confirmed cases; the allocation was redrawn in every bootstrap replicate, and the model was fitted across ten imputed allocations, so that the intervals include the uncertainty in where these deaths occurred. A sensitivity analysis of the decomposition removes these weeks instead. Five weeks of 2022 survive only as a single total derived from two cumulative reports; they count towards year totals but are treated as unobserved in weekly analyses.

*Years included.* 2017 was excluded from inference before analysis, because 18 of its weeks have no report, including the opening and peak weeks of the transmission season. 2020 and 2021, which overlap the COVID-19 pandemic, are included as they are; changes in reported Lassa fever during the pandemic have been attributed both to under-reporting and to reduced transmission[2,3]. The analysis scripts were written and tested on a preliminary extract before the final data were analysed.

*Deviations from the plan.* Four dated addenda were added after the plan was fixed[4]. Three (5 October 2026), made after the first results on the final data, added the comparison of printed cumulative with summed weekly figures (Addendum 1), the check against the Weekly Epidemiological Report (Addendum 2) and the comparisons on the printed series (Addendum 3). The fourth (9 October 2026) records that the extraction checks for 2021-2023, completed after the plan was fixed, identified 30 values entered incorrectly; the extract was corrected and every analysis rerun on it before submission. No analysis, estimand or data-handling rule changed, and results for 2025-2026 were unaffected.

*Roles of the analyses.* The primary analyses each answer a direct question about the reported change and make few assumptions beyond the counts. The roles were fixed before analysis on grounds of design and of what the data can support: the model cannot separate a national change in ascertainment from a national change in care, and isolated detection cannot separate testing prompted by a death from retrospective death investigation and speaks mainly to states with few cases. Within each analysis, other outputs are reported as secondary, sensitivity or descriptive.

*Phase-matched comparison.* Later percentiles than the 50th were not used, because a late percentile of a truncated total describes the truncation as much as the year.

*Decomposition.* A split with every state separate is a secondary, noisy output. Cutting every year to week 36 makes each pair compare the same weeks; transmission runs from about December to April, so the later weeks hold the inter-epidemic troughs and the start of the next transmission season. Two sensitivity analyses repeat the split with weeks of nationally reported deaths removed rather than allocated, and with full years.

*Tipping point.* The same calculation on the reconstruction from weekly reports, and against a pooled earlier reference, is secondary.

*Common or localised change (secondary).* I fitted a hierarchical binomial model of deaths among confirmed cases with random intercepts for year, state, and state within year[5], using brms[6]. With one row per state-year, the state-within-year term also carries overdispersion. If CFR moves between years for a system-wide reason such as ascertainment, variation between years should dominate; if it moves because of local change, variation between states within a year should. A national change in care would also appear as common variation, so the pattern does not identify a cause. The main estimand is the posterior probability that the year standard deviation exceeds the state-within-year standard deviation, reported without a threshold. The state-within-year term also absorbs data-quality noise, which tilts this comparison towards a local explanation. From the same fit I report the 2025-2026 contrast in year effects and the 2026 deviations of the states that stand alone in that pair. As in the decomposition, every year was cut to week 36. Sensitivity analyses used a beta-binomial likelihood, wider and tighter priors on the standard deviations, refits without Edo and without Ondo, which hold most confirmed cases, and full years.

*Detection triggered by death (secondary).* The reports do not say how many confirmed cases were first sampled at or after death. As a proxy, I defined an isolated detection as a state-week with confirmed cases after three weeks in which that state confirmed none, and asked whether a death was reported in that week or the next. If detection were blind to severity, a window holding $n$ confirmed cases would include a death with probability $1 - (1 - p)\text{\^{}}n$. The main estimand is the ratio of the observed to the expected share with $p$ each state's own CFR, estimated from its weeks outside the event windows and pooled over 2018-2026. Each state's CFR is itself an estimate, often from few cases, so every bootstrap replicate redraws it from a Jeffreys beta distribution[7]; the interval therefore reflects uncertainty in the expected share as well as in the observed one. Design simulations showed that a single national CFR cannot separate detection triggered by death from states differing in CFR. Using each state's own CFR removes that confusion, at the cost of detecting only substantial detection bias. The year's national CFR and the CFR among patients admitted to a specialist treatment centre in the LASCOPE cohort are used as secondary nulls[8]; the LASCOPE CFR is a reference point rather than a bound, since treatment may act to lower it and the exclusion of mild cases raises it. The windows cannot be estimated from these data, because the interval from case to death is not reported; they were taken from LASCOPE, in which most deaths occurred within a week of admission, and are varied in a sensitivity analysis because those timings describe patients who reached specialist care. Events are also compared by how often the state reported suspected cases that year: in at least half the weeks for which suspected cases are given by state, or fewer. A week with no suspected cases cannot be told apart from one in which cases were not recognised or reported, so this describes reporting, not surveillance capacity.

*Descriptive analyses.* The proportion of suspected cases confirmed, nationally and by state; use of the probable category by state and year; and the cumulative figures printed in each year's final report against the sum of that year's weekly reports.

*Software.* Analyses used R 4.6.1[9], with the model fitted in brms 2.23.0[6] through cmdstanr 0.9.0[10].

*Uncertainty.* The bootstrap resamples state-weeks, as clusters of cases, within each year[11]. Resampling state-weeks independently ignores correlation between a state's consecutive weeks, so the bootstrap intervals are probably somewhat too narrow. Intervals for isolated detection come from resampling state-years. Model uncertainty is the posterior across ten imputed datasets.

### Supplementary results

*Definitions.* The confirmed-case definition is worded identically in every report from 2017 week 5 to 2026 week 36. Only the probable-case definition changed: from 2019 week 25 it covers a suspected case "who died or absconded without collection of specimen", where earlier reports covered only those who died. Confirmed cases were reported from 19 states in 2025 and 22 in 2026.

*Decomposition.* With deaths reported only nationally removed rather than allocated, components changed by at most +1.15 percentage points. With full years instead of years cut to week 36, the 2025-2026 within-state component was -2.2 percentage points and the between-state component +1.3 percentage points. With every state separate, the largest within-state contributions came from Bauchi, where the CFR went from 8.2% to 13.6% (+1.3 percentage points, 95% CI -0.2 to 2.8), and Edo (+0.9 percentage points, 95% CI -0.4 to 2.3); the largest between-state contribution came from the growth of Benue's cases, from 15 to 61 (+0.6 percentage points).

*Tipping point.* The published-scale number is roughly one in 16 of the people suspected of Lassa fever in 2026 who were not confirmed by week 23. Against the pooled earlier years it would take 105 (95% CI 34 to 178).

*Common or localised change.* The 2025-2026 contrast in year effects was -0.22 on the logit scale (95% CrI -0.74 to 0.23), with a posterior probability of 0.17 of being above zero (Figure S1). Across the sensitivity fits the probability that the year standard deviation exceeded the state-within-year standard deviation ranged from 0.11 to 0.71; without Edo it was 0.14 and without Ondo 0.28. The answer depended on how overdispersion was modelled. The year standard deviation was similar in every fit (medians 0.33 to 0.42), while the state-within-year standard deviation was 0.53 with the binomial likelihood and 0.24 with the beta-binomial, which carries overdispersion in a separate term.

*Detection triggered by death.* I identified 336 isolated detections in 174 state-years. The ratio of observed to expected share with a death included one. Against the national CFR the ratio was 1.48 (95% CI 1.27 to 1.69), and against the LASCOPE CFR 1.97 (95% CI 1.67 to 2.26) (Figure S2). High-burden states report almost continuously and contribute few events, so this result speaks mainly to low- and mid-burden states. By stratum, the ratio was 0.99 (95% CI 0.74 to 1.24) in states that stand alone in the decomposition and 1.32 (95% CI 0.89 to 1.52) in the rest, 1.25 (95% CI 0.87 to 1.45) in weeks 1-13 and 1.10 (95% CI 0.80 to 1.30) later in the year, and 0.96 (95% CI 0.73 to 1.20) in state-years that reported suspected cases in at least half their weeks and 1.23 (95% CI 0.72 to 1.47) in the rest; every interval included one. The windows come from patients who reached a specialist treatment centre[8], and the interval from confirmation to death may differ elsewhere.

*State records.* Analyses of state and treatment-centre records give counts that differ from the reports for the same years: Ebonyi's records hold 368 confirmed cases and 125 deaths for 2018-2023[12], against 313 and 99 in the weekly reports, and the Edo treatment centre, which also takes referrals from neighbouring states, confirmed 652 cases in 2023[13], against 351 reported for Edo. In Ondo, about a quarter of suspected cases with a test result were positive in each year from 2017 to 2020, falling to 18% in 2021[14], the year in which the national proportion of suspected cases confirmed also fell.

**Table S1.** Checks of the extraction against the totals printed in each report, by year. Entry errors are extracted values found to differ from the report, all corrected before the final run. Inconsistent weeks are those in which the report's page-one totals disagree with its own state table; the state table was kept.

| Year | Check | Reports | Values extracted | Entry errors | Error rate (%) | Inconsistent weeks |
|---|---|---|---|---|---|---|
| 2017 | not checked | 34 | 558 | - | - | - |
| 2018 | Highlights text, state by state (confirmed, deaths) | 51 | 360 | 5 | 1.4 | 0 |
| 2019 | Highlights text, state by state (confirmed, deaths) | 50 | 426 | 8 | 1.9 | 0 |
| 2020 | page 1 Table 1 national totals | 53 | 2,306 | 14 | 0.6 | 1 |
| 2021 | page 1 Table 1 national totals; flagged weeks read against Table 3 | 47 | 1,580 | 7 | 0.4 | 6 |
| 2022 | Table 1 national totals; damaged text layer read against Table 3 Totals | 46 | 2,436 | 11 | 0.4 | 0 |
| 2023 | page 1 Table 1 national totals; flagged weeks read against Table 3 | 51 | 2,948 | 13 | 0.4 | 3 |
| 2024 | Table 3 printed Totals, every week | 51 | 3,324 | 12 | 0.4 | 2 |
| 2025 | Table 3 printed Totals, every week | 52 | 2,528 | 2 | 0.1 | 1 |
| 2026 | Table 3 printed Totals, every week | 36 | 2,244 | 8 | 0.4 | 2 |
| 2018-2026 | | 437 | 18,152 | 80 | 0.4 | 15 |

**Table S2.** RECORD checklist for observational studies using routinely collected health data. Items are abbreviated from the RECORD statement (Benchimol and colleagues, 2015; CC BY) and its parent STROBE items; see the statement for their full wording. Locations are sections of the main text and this appendix. NA, not applicable.

| Item | Recommendation | Location |
|---|---|---|
| 1a | Study design named in the title or abstract | Abstract, Methods |
| 1b | Informative, balanced summary in the abstract | Abstract |
| RECORD 1.1 | Type of data, and name of the database, in the title or abstract | Abstract, Methods (NCDC situation reports) |
| RECORD 1.2 | Geographic region and timeframe in the title or abstract | Title; Abstract |
| RECORD 1.3 | Linkage between databases stated in the title or abstract | NA: no linkage |
| 2 | Scientific background and rationale | Introduction |
| 3 | Objectives, including pre-specified hypotheses | Introduction, final paragraph; Methods, Data (pre-specified plan) |
| 4 | Key elements of the study design early in the paper | Methods, Data (first sentence) |
| 5 | Setting, locations and relevant dates | Methods, Data and Definitions |
| 6a | Sources and methods of selection of the population | Methods, Data; appendix, Extraction and data handling |
| 6b | Matching criteria | NA |
| RECORD 6.1 | Methods of population selection, such as codes or algorithms | Methods, Definitions; appendix, Supplementary results, Definitions (case definitions as printed) |
| RECORD 6.2 | Validation of the codes or algorithms used to select the population | Methods, Data (checks of the extraction); appendix, Extraction and data handling; Table S1 |
| RECORD 6.3 | Flow diagram of data linkage | NA: no linkage |
| 7 | Definitions of outcomes, exposures, predictors, confounders and effect modifiers | Methods, Definitions; Table 1 of the main text |
| RECORD 7.1 | Codes and algorithms used to classify exposures and outcomes | Methods, Definitions; appendix, Supplementary results, Definitions |

| Item | Recommendation | Location |
|---|---|---|
| 8 | Data sources and methods of measurement for each variable | Methods, Data; appendix, Extraction and data handling |
| 9 | Efforts to address potential sources of bias | Methods, Mechanisms and evaluability; Table 1 of the main text; Results, Evaluability of candidate mechanisms |
| 10 | How the study size was arrived at | Methods, Data (all reports to the data freeze); Results, opening paragraph |
| 11 | Handling of quantitative variables and groupings | Methods, Analyses (phase points; threshold of 30 confirmed cases) |
| 12a | Statistical methods | Methods, Analyses; appendix, Supplementary methods |
| 12b | Subgroups and interactions | Appendix, Supplementary methods (decomposition with every state; strata of isolated detection) |
| 12c | Missing data | Methods, Data; appendix, Extraction and data handling |
| 12d | Loss to follow-up, matching or sampling strategy | NA: aggregate counts; follow-up of outcomes discussed in Discussion |
| 12e | Sensitivity analyses | Appendix, Supplementary methods and results |
| RECORD 12.1 | Extent of the investigators' access to the database | Methods, Data (published reports only) |
| RECORD 12.2 | Data cleaning methods | Methods, Data; appendix, Extraction and data handling; Table S1 |
| RECORD 12.3 | Data linkage across databases | NA: no linkage; the Weekly Epidemiological Report was used as a check (Methods, Analyses) |
| 13 | Numbers at each stage of the study | Results, opening paragraph; appendix, Extraction and data handling |
| RECORD 13.1 | Selection of the population included, including filtering on data quality and availability | Results, opening paragraph; appendix, Years included, and Extraction and data handling |
| 14a | Characteristics of the study population | Results, opening paragraph; Figure 1 |
| 14b | Missing data for each variable | Results, opening paragraph; appendix, Extraction and data handling |
| 14c | Follow-up time | NA |
| 15 | Numbers of outcome events or summary measures | Results; Figure 1 |
| 16a | Unadjusted and adjusted estimates with their precision | Results (95% CIs and CrIs); no adjustment for confounders, as the analysis is a decomposition of reported counts |
| 16b | Category boundaries for categorised continuous variables | Methods, Analyses |
| 16c | Relative risk translated into absolute risk | NA |
| 17 | Other analyses: subgroups, interactions, sensitivity analyses | Results, Secondary analyses; appendix, Supplementary results |
| 18 | Key results with reference to the objectives | Discussion, first paragraph |
| 19 | Limitations, including the direction and magnitude of potential bias | Discussion, limitations paragraph |
| RECORD 19.1 | Implications of using data not collected for the research question | Introduction (reports built for situational awareness); Discussion, limitations paragraph and paragraphs on the two counts of deaths and strain on the system |
| 20 | Cautious overall interpretation | Discussion |
| 21 | Generalisability | Discussion, paragraph on other endemic zoonoses |
| 22 | Funding and the role of the funders | Abstract, Funding; Methods, Role of the funding source |

| Item | Recommendation | Location |
|---|---|---|
| RECORD 22.1 | Access to the protocol, raw data and programming code | Data sharing |

### Supplementary figures

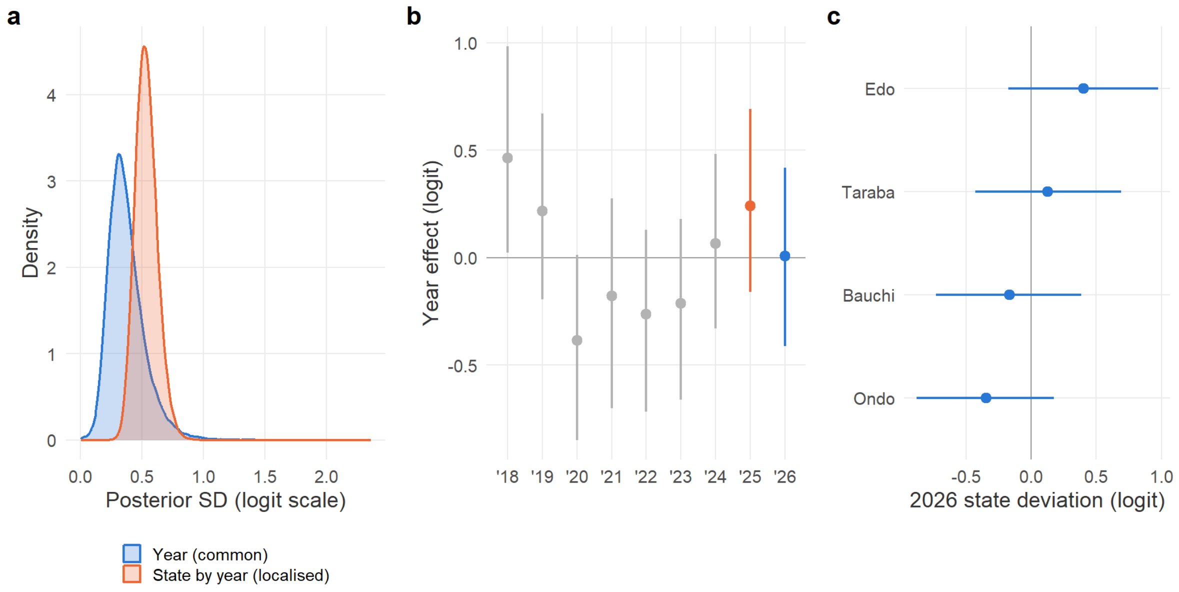

Figure S1: Common or localised change, a secondary analysis. (a) Posterior standard deviations of the year and state-within-year terms. (b) Year effects. (c) 2026 deviations for the states in the 2025-2026 roster.

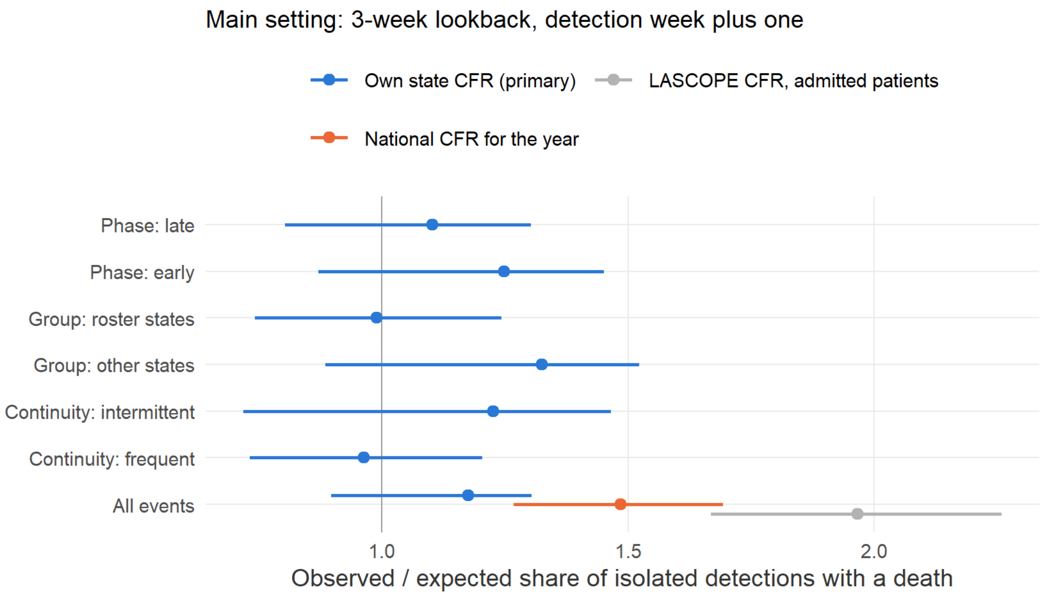

Figure S2: Detection triggered by death, a secondary analysis. Isolated detections with a death, observed against expected under severity-blind detection, with the national-CFR and LASCOPE nulls and by stratum.

### References

1 Benchimol EI, Smeeth L, Guttmann A, *et al.* The REporting of studies Conducted using Observational Routinely-collected health Data (RECORD) statement. *PLoS Medicine* 2015; **12**: e1001885.

2 Musa SS, Zhao S, Abdullahi ZU, Habib AG, He D. COVID-19 and Lassa fever in Nigeria: A deadly alliance? *International Journal of Infectious Diseases* 2022; **117**: 45–7.

3 Reuben RC, Gyar SD, Makut MD, Adoga MP. Co-epidemics: Have measures against COVID-19 helped to reduce Lassa fever cases in Nigeria? *New Microbes and New Infections* 2021; **40**: 100851.

4 Simons D. Reported lassa fever case fatality in nigeria, 2017-2026: Pre-specified analysis plan and code, version 1.0. 2026. DOI:10.5281/zenodo.23135368.

5 Gelman A, Hill J. Data analysis using regression and multilevel/hierarchical models. Cambridge: Cambridge University Press, 2006.

6 Bürkner P-C. Brms: An R package for Bayesian multilevel models using Stan. *Journal of Statistical Software* 2017; **80**. DOI:10.18637/jss.v080.i01.

7 Brown LD, Cai TT, DasGupta A. Interval estimation for a binomial proportion. *Statistical Science* 2001; **16**: 101–33.

8 Duvignaud A, Jaspard M, Etafo IC, *et al.* Lassa fever outcomes and prognostic factors in Nigeria (LASCOPE): A prospective cohort study. *The Lancet Global Health* 2021; **9**: e469–78.

9 R Core Team. R: A language and environment for statistical computing. Vienna, Austria: R Foundation for Statistical Computing, 2026 https://www.R-project.org/.

10 Gabry J, Češnovar R, Johnson A, Bronder S. Cmdstanr: R interface to CmdStan. 2025 https://mc-stan.org/cmdstanr/.

11 Davison AC, Hinkley DV. Bootstrap methods and their application. Cambridge: Cambridge University Press, 1997 DOI:10.1017/CBO9780511802843.

12 Iziomo PM, Awosanya E, Taiwo OJ, *et al.* Temporal trends and epidemiological patterns of Lassa fever outbreaks in Ebonyi State, Nigeria: A retrospective study (2018-2023). *PLOS Global Public Health* 2026; **6**: e0006871.

13 Cadmus S, Awosanya E, Iziomo PM, *et al.* Deciphering the dynamics of Lassa fever outbreak in Edo State, Nigeria: A 3-year study of the epidemiologic patterns, severity profiles, and clinical management. *IJID One Health* 2025; **7**: 100059.

14 Cadmus S, Akinseye V, Cadmus E, *et al.* Drivers of Lassa fever in an endemic area of southwestern Nigeria (2017-2021): An epidemiological study. *PLoS One* 2025; **20**: e0327143.
